# RCT-based Social Return on Investment (SROI) of a home exercise programme for people with early dementia comparing in-person and blended delivery before and during the COVID-19 pandemic

**DOI:** 10.1101/2023.08.25.23294408

**Authors:** Kodchawan Doungsong, Ned Hartfiel, John Gladman, Rowan H Harwood, Rhiannon Tudor Edwards

## Abstract

**Background:** Regular exercise and community engagement may slow the rate of function loss for people with dementia. However, the evidence is uncertain regarding the cost-effectiveness and social return on investment (SROI) of home exercise with community referral for people with dementia. This study aimed to compare the social value generated from the in-person PrAISED programme delivered before March 2020 with a blended PrAISED programme delivered after March 2020.

**Methods:** SROI analysis was conducted alongside a randomised controlled trial (RCT). Of 205 patient participants and their carers who completed cost data, 61 completed an in-person programme before March 2020. Due to COVID-19 pandemic restrictions, 144 patient participants completed a blended programme consisting of a combination of in-person visits, phone calls and video conferencing with multidisciplinary team (MDT) members.

SROI analysis compared in-person and blended delivery formats. Five relevant and material outcomes were identified: three outcomes for patient participants (fear of falling, health-related quality of life, and social connection); one outcome for carer participants (carer strain index), and one outcome for the NHS (health service resource use). Data were collected at baseline and a 12-month follow-up.

**Results:** The in-person PrAISED programme generated SROI ratios ranging from £0.58 to £2.33 for every £1 invested. In-person PrAISED patient participants gained social value from improved health-related quality of life, social connection, and less fear of falling. In-person PrAISED carer participants acquired social value from less carer strain. The NHS gained benefit from less health care service resource use. However, the blended PrAISED programme generated lower SROI ratios ranging from a negative ratio to £0.08: £1.

**Conclusion:** Compared with the blended programme, the PrAISED in-person programme generated higher SROI ratios for people with early dementia. During the COVID-19 pandemic and its restrictions, a blended delivery of the programme and the curtailment of community activities resulted in lower SROI ratios during this period. An in-person PrAISED intervention with community referral is likely to provide better value for money than a blended one with limited community referral, despite the greater costs of the former.

**Trial registration:** ISRCTN15320670

## Introduction

Dementia is a complex and progressive syndrome causing cognitive impairment and restricted daily function (CDC, 2019; NICE, 2018). It affects the mental, physical and financial wellbeing of people living with dementia and their families (Vernooij-Dassen et al., 2021). In 2019, the World Health Organisation (WHO) reported that the global cost of dementia was $1.3 trillion and is expected to rise to $1.7 trillion by 2030 (WHO, 2021).

Preventing or delaying the onset of dementia by two years can have significant economic and societal benefits (Rakesh et al., 2017). For people with dementia, the ability to perform daily activities deteriorates as the disease progresses (Goldberg et al., 2019). For people with dementia and their carers, maintaining independence is a priority (Peach et al., 2017). Promoting regular exercise and engaging in community activities may slow the rate of functional loss (Forbes et al., 2015; Pitkälä et al., 2013; Roth, 2022). However, the cost-effectiveness of professionally supervised regular exercise for people with dementia is conflicting. One study indicated that a structured exercise programme over 12 months was not able to maintain cognitive function for people with dementia, and therefore was not cost-effective (Khan et al., 2019). Another study found that exercise therapy for people with dementia was not cost-effective in terms of quality adjusted life year (QALY) gains, but potentially cost effective when improvements in behavioural and psychological symptoms were considered (D’Amico et al., 2016). A home-based exercise programme for people with dementia in Finland reported reductions in hospital admissions and overall costs (Pitkälä et al., 2013) and another study demonstrated that 10 home-based sessions of community occupational therapy for people with dementia and their carers could be cost-effective (Graff et al., 2008).

PrAISED (promoting activity, independence, and stability in early dementia) was a multi-centre, pragmatic, randomised controlled trial (RCT), which was conducted between October 2018 and September 2022 (Bajwa et al., 2019; Harwood et al., 2023). Patient participants were recruited from five locations in England and were randomised to a control group (usual care) or an intervention group (PrAISED). The intervention was a specifically designed rehabilitation programme that provided a home-based exercise programme along with promoting access to community activities for people with early dementia (Booth et al., 2018; Logan et al., 2022). It was a complex intervention consisting of balance and strength building exercises, dual-task and functional activities training, and gait re-education. The PrAISED programme included home visits from a multidisciplinary team (MDT) of physiotherapists (PTs), occupational therapists (OTs) and rehabilitation support workers (RSWs) who encouraged patients to engage in 180 minutes of physical activity per week. Patient participants in the PrAISED group were offered up to 50 therapy sessions over 12 months while those in the usual care group received an assessment modelled on usual falls prevention care which consisted of an initial visit and up to two further visits (Harwood et al., 2023).

From October 2018 to February 2020, 61 patient participants completed the in-person PrAISED programme prior to the COVID-19 pandemic. Due to pandemic restrictions between March 2020 and September 2022, 144 patient participants completed a blended PrAISED programme which consisted of a combination of in-person visits, phone calls and video conferencing with MDT members. For blended programme patient participants, referral to and uptake of community activities was significantly reduced due to pandemic restrictions.

Providing a home-based exercise programme for people with dementia is a complex intervention. Recent guidelines from the Medical Research Council (MRC) suggest using a broad framework for economic evaluation to capture wide and diverse outcomes for complex interventions. In addition to cost-effectiveness analysis, the MRC recommends cost consequence analysis and cost-benefit analysis (CBA) (Skivington et al., 2021). Social return on investment (SROI), a type of social cost-benefit analysis, is becoming more commonly used to evaluate complex interventions (NEF, 2012; Skivington et al., 2021). SROI quantifies and values multiple outcomes to achieve a broader perspective than traditional cost-effectiveness analysis, which tends to focus on only one outcome (i.e., quality-adjusted life-year, QALY) (Nicholls et al., 2012; Bryan et al., 2007). SROI is a pragmatic form of social CBA, which is recommended in His Majesty’s (HM) Treasury Green Book in the United Kingdom (UK) to assess interventions that impact social welfare (HM Treasury, 2022; NEF, 2012). The framework for SROI is described in the Cabinet Office ‘A Guide to Social Return on Investment’ (Nicholls et al., 2012). SROI identifies and quantifies outcomes that are relevant to stakeholders and then determines financial proxies for these outcomes, which often do not have market values. This enables SROI ratios to be presented in monetary units (£). SROI adopts an individual approach to measuring the social value of outcomes rather than averaging across an entire intervention arm (Nicholls et al., 2012; Trotter et al., 2014). SROI can help inform decision-making by monetising and reporting the social value generated by a health or social care intervention.

There are few published SROI studies of interventions to improve the health and wellbeing of people with dementia. Previous studies include the impact of peer support groups, (Semple et al., 2015), art activities (Jones et al., 2020), and home exercise and community referral (Hartfiel et al., 2022). Our study intended to build upon the PrAISED feasibility study of home exercise and community referral for people with early dementia (Hartfiel et al., 2022). However, the impact of the COVID-19 pandemic rendered it impossible to conduct the PrAISED programme in-person, necessitating a change in the mode of delivery. Therefore, the aim of this SROI analysis is to compare the social value generated from the in-person PrAISED programme delivered before March 2020 with a blended PrAISED programme delivered after March 2020.

## Methods

The study design for the PrAISED multi-centre RCT has been previously reported (Harwood et al., 2023), covering important details such as the randomization method, participant selection criteria, informed consent, and ethical committee approval.

In this evaluation, a range of SROI ratios were generated using quantitative data collected during the PrAISED trial. SROI methodology contains the following steps: identifying stakeholders, developing a logic model, evidencing outcomes, valuing outcomes, calculating costs, and estimating the SROI ratio. This analysis was conducted in alignment with the 21-item SROI Quality Assessment Framework Tool (Hutchinson et al., 2018) (Appendix 1).

Key stakeholders were the people and organisations most affected by the PrAISED programme: patient participants, informal carer participants and the National Health Service (NHS). It was expected that patient participants would benefit from home exercise and community referral; carer participants would benefit from additional support provided by MDT home visits, and the NHS would benefit from less frequent health service use from PrAISED patient participants.

The logic model of the PrAISED intervention illustrates how the intervention intended to improve quality of life, lessen fear of falling, increase social connection, reduce carer strain and decrease health service resource use (Figure 1).

**Figure 1.**
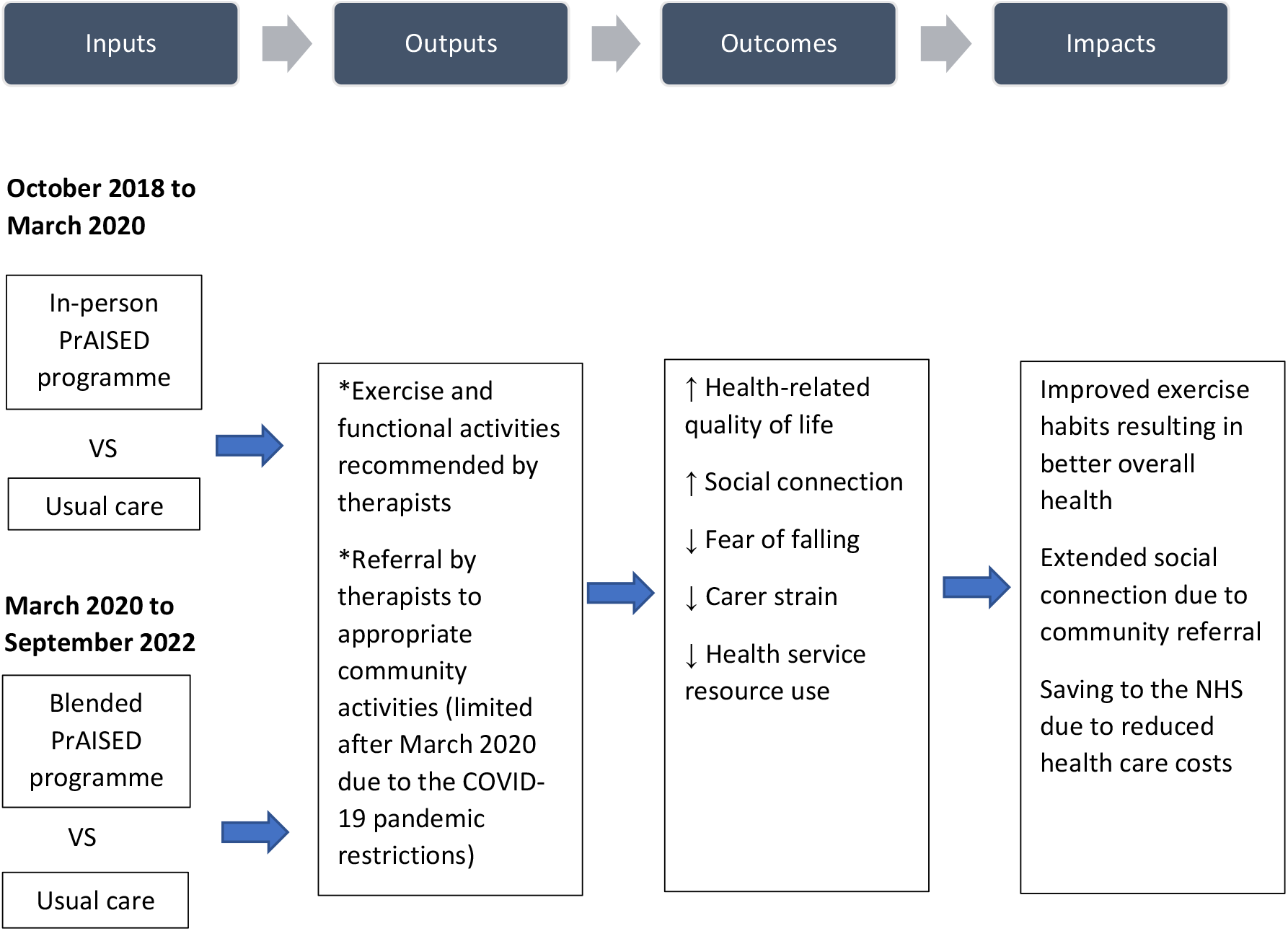
Logic model.

### Outcomes

Five relevant and material outcomes were identified from the logic model: improved health-related quality of life, increased social connection, less fear of falling, decreased carer strain, and reduced health service resource use. Four of these outcomes were measured and valued using baseline and follow-up questionnaires: fear of falling (Falls Efficacy Scale – International; FES-I) (Kempen et al., 2008), health-related quality of life (EQ5D-5L) (Herdman et al., 2011), carer strain (Carer Strain Index; CSI) (Robinson, 1983), and health service resource use (adapted Client Service Receipt Inventory (CSRI) (Table 1). Social connection was measured and valued using an adapted community activities questionnaire (CAQ) at follow-up (Appendix 5). Three of the outcome measures FES-I, EQ5D-5L, and CSI were validated scales frequently used in studies involving persons with dementia.

**Table 1.** Outcome measures for the three key stakeholders.

| Stakeholder | Outcome | Outcome measure | Completed by | Source of outcome values | Outcome values |
| --- | --- | --- | --- | --- | --- |
| Patient participant | High confidence (less fear of falling) | Falls Efficacy scale - International (FES-I) | Patient participant | SVB | £13,080 per person per year |
| Patient participant | Good overall health (increased health-related quality of life) | Euroqol EQ5D-5L | Carer participant | SVB | £20,141 per person per year |
| Patient participant | Regular participation in a social group (More social connection) | Community Activities Questionnaire (CAQ) | Carer participant | SVB | £1,850 per person per year |
| Carer participant | Able to rely on family (Less carer strain) | Carer Strain Index (CSI) | Carer participant | SVB | £6,784 per person per year |
| NHS | Health service resource use | Client Service Receipt Inventory (CSRI) | Carer participant | Financial savings from difference in health service resource use at baseline and follow-up |  |

FES-I measured whether home exercise could generate more confidence in patient participants performing activities of daily living and therefore reduce the risk of falls. Health-related quality of life as measured by the EQ5D is recommended by the National Institute for Health and Care Excellence (NICE) for conducting economic evaluations of complex public health interventions (NICE, 2019). The CAQ measured the degree to which patients increased their engagement in community activities due to the PrAISED intervention. For carer participants, the CSI was a key outcome measure, assessing whether PrAISED lessened the burden of care on family members or carers. Finally, the CSRI form measured whether PrAISED resulted in reduced healthcare service use by patient participants in comparison with usual care (Table 1).

For four outcomes (FES-I, EQ5D-5L, CAQ, and CSI), financial proxies were assigned from the Social Value Bank (SVB). For three outcomes (FES-I, EQ5D-5L, and CSI), financial proxies were assigned using two cut-off points for participants who improved by ≥ 5%, and ≥ 10%. For CAQ, a financial proxy was assigned for those participants who participated in community activities at least once a month (HACT, 2018). The SVB is a databank of methodologically consistent unit costs for outcome indicators. It is based on ‘wellbeing valuation’ and recommended in the HM Treasury Green Book as a robust method for financial appraisal and evaluation (Trotter et al., 2014). Monetised values from the SVB included £13,080 for high confidence (less fear of falling); £20,141 for good overall health (health-related quality of life); £1,850 for regular attendance at a social group (social connection); and £6,784 for being able to rely on family (less carer strain) (HACT, 2018).

### Health Service Resource Use

Outcomes for the NHS were assessed by measuring the health service resource use of patient participants during the previous three months before the baseline and 12-month follow-up. Cost data were obtained from NHS unit costs and Personal Social Services Research Unit 2020/2021 (Jones & Burns, 2021; NHS, 2022) (Appendix 4). Health service resource use included contact with general practitioners (GPs), nurses, physiotherapists and other hospital services (Appendix 4). Health service resource use analysis compared PrAISED with usual care patient participants for both in-person and blended delivery formats.

### Training and Delivery Costs

Total costs for the PrAISED intervention included the following categories: MDT training costs, MDT staffing and transport costs, instructional materials and exercise equipment for patient participants. Total costs per patient participant were calculated for both in-person and blended formats.

### SROI ratios

After outcomes and costs were monetised, SROI ratios were calculated by dividing the social value per participant by the cost per participant. Intervention participants were compared with usual care participants for both the in-person and blended formats. In most SROI evaluations, deadweight, attribution, displacement, and drop-off are considered (Nicholls et al., 2012). However, our study design was an RCT, and therefore it was unnecessary to calculate these factors. Discounting was also unnecessary because all costs and outcomes were measured within 12-months of the baseline assessment.

## Results

Of the 365 trial participants, 75 (21%) did not complete 12-month follow-up outcome data and an additional 85 (23%) did not complete cost data. Therefore, 56% (205/365) completed cost data at both baseline and follow-up. Of these, 61 participants completed the in-person programme, and 144 participants completed a blended programme. For blended PrAISED participants, approximately 69% of sessions were received in-person and 31% were experienced by telephone or videoconference.

Baseline characteristics were similar between in-person and blended patient participants. The mean age was 81 and 79 years for in-person and blended programmes, respectively. Most patient participants in both programmes were male, married, and white ethnic (Appendix 2).

### In-person programme

SROI results for the in-person programme showed that when compared with usual care, a higher proportion of in-person PrAISED patient participants reported improvements of ≥5% in fear of falling, social connection, and carer strain (Table 2). Social value was also higher for in-person PrAISED patient and carer participants when compared with usual care at both the 5% and 10% cut-off points (Table 3).

**Table 2.** Quantity of relevant outcomes measures for in-person and blended programmes.

| Outcome measure | Programme | Complete cases | Group | Improvement of $\geq 5\%$ | Improvement of $\geq 10\%$ |
| --- | --- | --- | --- | --- | --- |
| FES-I<sup>1</sup> | In-person | n=59 | PrAISED (n=29) | 7/29 (24%) | 7/29 (24%) |
|  |  |  | Usual care (n=30) | 4/30 (13%) | 4/30 (13%) |
|  | Blended | n=127 | PrAISED (n=69) | 10/69 (14%) | 10/69 (14%) |
|  |  |  | Usual care (n=58) | 15/58 (26%) | 15/58 (26%) |
| EQ5D-5L<sup>2</sup> | In-person | n=60 | PrAISED (n=30) | 6/30 (20%) | 6/30 (20%) |
|  |  |  | Usual care (n=30) | 9/30 (30%) | 4/30 (13%) |
|  | Blended | n=141 | PrAISED (n=79) | 16/79 (22%) | 12/79 (15%) |
|  |  |  | Usual care (n=62) | 10/62 (16%) | 5/62 (8%) |
| CAQ<sup>3</sup> | In-person | n=60 | PrAISED (n=30) | 20/30 (67%) | 20/30 (67%) |
|  |  |  | Usual care (n=30) | 14/30 (47%) | 14/30 (47%) |
|  | Blended | n=141 | PrAISED (n=78) | 19/78 (24%) | 19/78 (24%) |
|  |  |  | Usual care (n=63) | 11/63 (17%) | 11/63 (17%) |
| CSI<sup>4</sup> | In-person | n=60 | PrAISED (n=31) | 14/31 (45%) | 9/31 (29%) |
|  |  |  | Usual care (n=29) | 8/29 (28%) | 5/29 (17%) |
|  | Blended | n=130 | PrAISED (n=70) | 24/70(34%) | 18/70 (25%) |
|  |  |  | Usual care (n=60) | 26/60 (43%) | 15/60 (25%) |

<sup>1</sup> Falls Efficacy scale – International; <sup>2</sup> Euroqol EQ5D-5L; <sup>3</sup>Community activities questionnaire; <sup>4</sup> Carer Strain Index

**Table 3.** Valuing Outcomes for In-person Programme.

| Group | Outcome measure | Value from Social Value Bank | Quantity improved by ≥ 5% | Total Social Value | Social value per participant | Quantity improved by ≥10% | Total Social Value | Social value per participant |
| --- | --- | --- | --- | --- | --- | --- | --- | --- |
| PrAISED | FES-I | £13,080 per year - high confidence | 7/29 (24%) | £91,560 | £3,157 | 7/29 (24%) | £91,560 | £3,157 |
| Usual care | FES-I | £13,080 per year - high confidence | 4/30 (13%) | £52,320 | £1,744 | 4/30 (13%) | £52,320 | £1,744 |
| PrAISED | EQ5D-5L | £20,141 per year – good overall health | 6/30 (20%) | £120,846 | £4,028 | 6/30 (20%) | £120,846 | £4,028 |
| Usual care | EQ5D-5L | £20,141 per year – good overall health | 9/30 (30%) | £181,269 | £6,042 | 4/30 (13%) | £80,564 | £2,685 |
| PrAISED | CAQ | £1,850 per year - regular attendance in social group | 20/30 (67%) | £37,000 | £1,233 | 20/30 (67%) | £37,000 | £1,233 |
| Usual care | CAQ | £1,850 per year - regular attendance in social group | 14/30 (47%) | £25,900 | £863 | 14/30 (47%) | £25,900 | £863 |
| PrAISED | CSI | £6,784 per year - able to rely on family | 14/31 (45%) | £94,676 | £3,064 | 9/31 (29%) | £61,056 | £1,970 |
| Usual care | CSI | £6,784 per year - able to rely on family | 8/29 (28%) | £54,272 | £1,871 | 5/29 (17%) | £33,920 | £1,170 |
| Outcome 1: Less fear of falling - difference between groups |  |  |  |  | £1,413 |  |  | £1,413 |
| Outcome 2: Health related quality of life - difference between groups |  |  |  |  | -£2,014 |  |  | £1,343 |
| Outcome 3: Social connection - difference between groups |  |  |  |  | £370 |  |  | £370 |
| Outcome 4: Carer strain - difference between groups |  |  |  |  | £1,193 |  |  | £800 |
| **Total social value per participant** |  |  |  |  | **£962** |  |  | **£3,926** |

### Blended programme

SROI results for the blended programme indicated that when compared with usual care, a lower proportion of blended PrAISED patient participants reported improvements of ≥5% in fear of falling and carer strain (Table 2). Social value was also lower for blended PrAISED patient participants and only slightly higher for blended PrAISED carer participants when compared with usual care at both the 5% and 10% cut-off points (Table 4).

**Table 4.** Valuing Outcomes for Blended Programme.

| Group | Outcome measure | Value from Social Value Bank | Quantity improved by $\geq 5\%$ | Total Social Value | Social value per participant | Quantity improved by $\geq 10\%$ | Total Social Value | Social value per participant |
| --- | --- | --- | --- | --- | --- | --- | --- | --- |
| PrAISED | FES-I | £13,080 per year - high confidence | 10/69 (14%) | £130,800 | £1,896 | 10/69 (17%) | £130,800 | £1,896 |
| Usual care | FES-I | £13,080 per year - high confidence | 15/58 (26%) | £196,200 | £3,383 | 15/58 (26%) | £196,200 | £3,383 |
| PrAISED | EQ5D-5L | £20,141 per year – good overall health | 16/79 (22%) | £322,256 | £4,079 | 12/79 (15%) | £241,692 | £3,059 |
| Usual care | EQ5D-5L | £20,141 per year – good overall health | 10/62 (16%) | £201,410 | £3,249 | 5/62 (8%) | £100,705 | £1,624 |
| PrAISED | CAQ | £1,850 per year - regular attendance in social group | 19/78 (24%) | £35,150 | £451 | 19/78 (24%) | £35,150 | £451 |
| Usual care | CAQ | £1,850 per year - regular attendance in social group | 11/63 (17%) | £20,350 | £323 | 11/63 (17%) | £20,350 | £323 |
| PrAISED | CSI | £6,784 per year - able to rely on family | 24/70(34%) | £162,816 | £2,326 | 18/70 (26%) | £122,112 | £1,744 |
| Usual care | CSI | £6,784 per year - able to rely on family | 26/60 (43%) | £176,384 | £2,940 | 15/60 (25%) | £101,760 | £1,696 |
| Outcome 1: Less fear of falling - difference between groups |  |  |  |  | -£1,487 |  |  | -£1,487 |
| Outcome 2: Health related quality of life - difference between groups |  |  |  |  | £830 |  |  | £1,435 |
| Outcome 3: Social connection - difference between groups |  |  |  |  | £128 |  |  | £128 |
| Outcome 4: Carer strain - difference between groups |  |  |  |  | -£614 |  |  | £48 |
| **Total social value per participant** |  |  |  |  | **-£1,143** |  |  | **£124** |

**Table 5.** SROI ratios.

| Category | Quantify by improved $\geq 5\%$ | | Quantify by improved $\geq 10\%$ | |
| --- | --- | --- | --- | --- |
|  | In-Person | Blended | In-Person | Blended |
| Outcome 1 - improved confidence (less fear of falling) | £1,413 | -£1,487 | £1,413 | -£1,487 |
| Outcome 2 - improved health related quality of life | -£2,014 | £830 | £1,343 | £1,435 |
| Outcome 3 - increased social connection | £370 | £128 | £370 | £128 |
| Outcome 4 - less carer strain | £1,193 | -£614 | £800 | £48 |
| NHS health service resource use | £11.71 | -£23.36 | £11.71 | -£23.36 |
| **Total social value for all stakeholders** | **£974** | **-£1,166** | **£3,938** | **£100** |
| **Total cost** | **£1,688** | **£1,244** | **£1,688** | **£1,244** |
| **SROI ratio** | **£0.58: £1** | **Negative ratio** | **£2.33: £1** | **£0.08: £1** |

### Training and delivery costs

The NHS was responsible for training and delivery costs of the 12-month PrAISED programme. Overall, 61 therapists were trained to deliver the PrAISED programme to 183 patient participants. In-person training costs averaged £184 per patient participant compared to £139 per patient participant for online training (Appendix 3).

The difference in delivery costs between PrAISED and usual care was £1,504 per participant for the in-person programme (Appendix 3), and £1,105 per participant for the blended programme. Total costs for the in-person programme were higher than blended programme by £444 per participant (Appendix 3).

### Health service resource use

For the in-person programme, health service resource use was £11.71 less per PrAISED patient participant in comparison with usual care, mostly due to PrAISED patient participants reporting a 20% reduction in outpatient visits during the 12-month intervention (Appendix 4). For the blended programme, health service resource use was £23.36 more per PrAISED patient participant in comparison with usual care, partly due to an 8% increase in outpatient visits by PrAISED patient participants during the 12-month intervention (Appendix 4).

### SROI ratios

SROI ratios comparing the in-person programme with usual care ranged from £0.58: £1 to £2.33: £1. In contrast, SROI ratios for the blended programme ranged from a negative ratio to £0.08:£1.

## Discussion

PrAISED was designed as an in-person exercise rehabilitation programme delivered in the homes of people with early dementia. Due to COVID-19, the intervention had to be adapted to a blended programme. A substantial difference was found in SROI ratios between the pre-COVID in-person delivery and the blended delivery during COVID. The in-person programme SROI ratios ranged from £0.58 to £2.33 for every £1 invested, while the SROI ratios for the blended programme ranged from a negative ratio to £0.08: £1.

The range of SROI ratios for the in-person PrAISED programme was lower than the SROI ratios from the in-person PrAISED feasibility trial, which ranged from £3.46 to £5.94 for every £1 invested (Hartfiel et al., 2022). Possible explanations for differences in results include variations in the population recruited, expertise of the clinicians delivering the programme, the small sample sizes, and different thresholds for assigning social values.

The variation in the population recruited refers to a higher proportion of feasibility study participants enrolling from the Join Dementia Research (JDR) service. These participants tended to have higher motivation and engaged more fully with the PrAISED programme which resulted in higher outcomes. The feasibility trial had a higher proportion of MDT members who were therapists within NHS Mental Health Services for Older People (MHSOP). These therapists had more training and experience in working with people with dementia, which may have also led to higher outcomes.

Differences in SROI ratios between the feasibility study and our study may have also been due to using different thresholds for assigning social value. In the feasibility trial, SVB financial proxies were assigned to participants who either ‘stayed the same’ or ‘improved’ between baseline and follow-up. However, one of the main principles in social value accounting is to ‘only include what is material’ and social value guidance states that ‘materiality is essentially a matter of professional judgment’ (Social Value International Organisation, 2022).

In our study, SVB financial proxies were assigned to those participants who reported improvements of ≥ 5% and ≥ 10%, which in our professional judgment may provide a more accurate measure of a material improvement.

The lower SROI ratios for the blended programme may have been due to restrictions on the in-person delivery during the pandemic, and the inability to refer patients to community activities. During the pandemic, community activities became unavailable or unattractive to older people with early dementia who may have considered themselves at high risk of COVID-19. Regular engagement with community groups is significantly associated with slower cognitive decline for people with dementia and lower risk of developing dementia (Fancourt et al., 2020; Roth, 2022). Telephone and video conference delivery made it more difficult for the MDT to develop therapeutic relationships with participants and to help participants advance toward goals that required their physical presence (di Lorito et al., 2021). Patient participants often engaged more deeply with PrAISED exercises when MDT members were physically present (di Lorito et al., 2021).

### Strengths of this study

To our knowledge, this SROI evaluation is the first study to compare the social value of in-person versus blended delivery of a home-based exercise programme to people with dementia. The RCT design helped ensure that social value was not overestimated, and the societal perspective using multiple outcomes helped ensure that important benefits were not overlooked. In addition, the majority of outcomes were based on quantitative data collected from both patient and carer participants at baseline and follow-up. Finally, the social value sets used in this study were derived from wellbeing valuation, a robust method recommended for measuring and valuing social CBA (HM Treasury, 2022).

### Limitations of this study

The main limitations were due to the COVID-19 pandemic, which could not have been anticipated. The sample sizes for the in-person and blended participants were unplanned. It was not possible to determine to what degree the change in the mode of delivery or the change in the social context during the pandemic explained the differences in SROI ratios between the in-person and blended phases.

Another limitation is that the Social Value Bank (SVB) does not have monetary proxies for every possible material outcome. Monetised values in this study were applied for outcomes closely associated with specific SVB values (HACT, 2018). However, matching study outcomes with specific SVB values depends on researcher discretion, which could lead to overestimating or underestimating social value.

## Conclusion

Compared with blended delivery, the PrAISED in-person exercise programme generated higher SROI ratios for people with early dementia. During the COVID-19 pandemic, a blended delivery of the exercise programme and the curtailment of community activities may explain why there was a negative SROI ratio during this period. These findings suggest that in-person delivery with community referral is likely to provide better value for money than a blended delivery with limited community referral, despite the greater costs of the former. The findings also illustrate how the social context in which the intervention was implemented could significantly impact the value for money.

## Data Availability

All data produced in the present work are contained in the manuscript

