## Supplementary file for "RCT-based Social Return on Investment (SROI) of a home exercise programme for people with early dementia comparing in-person and blended delivery before and during the COVID-19 pandemic"

#### Appendix 1 - SROI Quality Assessment Framework Tool

##### Appendix 1, Table 1

| Research Question | Scoring | Notes |
|---|---|---|
| Was a well-defined question posed? | Yes, p.2 | The study question comparing in-person and blended PrAISED programmes was explained in the Introduction section. |
| Reason for use of SROI Method | | |
| Were authors transparent about why SROI methodology was chosen? (e.g. strategic planning/funding requirements) | Yes, pp.1 – 2 | SROI methodology was chosen as part of health economic evaluation of PrAISED programme (Harwood et al., 2023). The reasons for choosing SROI methodology were discussed in Introduction and Methods section. |
| Did authors report relevant background literature/justify the need for the study? | Yes, pp.1 - 2 | Relevant background was provided in the Introduction section. |
| Scope | | |
| Was the range of stakeholders included/excluded justified? | Yes, pp.2 – 3 | Key stakeholders and reasons for inclusion were provided in the Methods section |
| Was the range of stakeholders wide enough to adequately answer the research question? (principle of understanding change) | Yes, pp.2 - 4 | The range of stakeholders was discussed in the Methods section. |
| Was it clear how stakeholders were involved and what data would be gathered from them? | Yes, pp.2 – 4 | The involvement of stakeholders, was provided in the Methods section. |
| Was ethics obtained/informed consent provided? | Yes, p.2 | Ethic approval was presented in the Method section |
| Theory of change/impact map | | |
| Was the theory of change clear? i.e. the relationships between inputs, outputs and outcomes | Yes, pp.2 - 3 | The logic model was presented in the Methods section |
| Are unintended outcomes (positive/negative) detailed? | Yes, pp.1 – 2 | Because dementia is a progressive disease, not all participants improved. This was discussed in the Introduction section. |
| Study Design | | |
| Was the study design appropriate for the study question? (Control group, pre-post) | Yes, pp.1 – 2 | This RCT study design was discussed in the Introduction. |
| Was the sample described in detail/was the sample justified? | Yes, p.1 | Sample sizes for before COVID and during COVID groups were justified in the Introduction section. |
| Analysis | | |
| Were inputs clear with non-monetized inputs valued appropriately? | Yes, see Supplementary File | The inputs (i.e., training and delivery costs) were clearly presented in Appendix 3. |
| Were capital costs, as well as operating costs included? | Yes, see Supplementary File | Capital costs for equipment and stationery were presented in Appendix 3. |
| Were costs that occur in the future 'discounted' to their present values? Was justification given for the discount rate used? | Yes, pp.2 – 5 | Discounting was discussed in the Methods section |
| Was dead-weight clearly described and calculated? | Yes, pp.2 - 5 | Deadweight was discussed in the Methods section |
| Were the indicators valid and comprehensive? (Were the sources of all values clearly identified?) | Yes, pp.3 – 4 | The indicators for noticeable outcomes and sources of financial value were discussed in the Method section. |
| Were the proxies valid and comprehensive? (Were the sources of all values clearly identified?) | Yes, pp.5 – 6 | Financial values were assigned from Social Value Bank. See Results section. |

| | | |
|---|---|---|
| Was length of benefit established and justified? (Drop-off) (In capital projects, did authors establish and differentiate between length of benefit and life expectancy of the asset?) | Yes, p.5 | The length of benefit was established at 12 months along with the duration of the intervention. See Results section. |
| Were limitations and biases reported? | Yes, p.12 | See Discussion section. |
| Was the final SROI ratio interpreted? | Yes, p.10 | The SROI ratios were interpreted in the Discussion section. |
| Was sensitivity analysis performed? Was justification provided for the range of values (or for key study parameters) in the sensitivity analysis? | Yes, p.4 | The range of SROI ratios were offered by comparing in-person and blended programmes for both cut-off points (≥5% and ≥10). See Methods section. |

Appendix 2 – Baseline characteristic

**Appendix 2, Table 1: Baseline characteristics of participants (complete cases n = 205)**

| | In-person programme (n=61) | | Blended programme (n=144) | |
|---|---|---|---|---|
| | PrAISED (n=30) | Usual care (n=31) | PrAISED (n=80) | Usual care (n=64) |
| Participant age (Mean) | 81 | 77 | 79 | 81 |
| Co-resident (Yes) | 25 (83%) | 28 (90%) | 66 (83%) | 48 (75%) |
| Gender (Male) | 19 (63%) | 19 (61%) | 45 (56%) | 37 (58%) |
| Marital status (Married) | 23 (77%) | 26 (84%) | 56 (70%) | 43 (67%) |
| Ethnic group (White) | 29 (97%) | 29 (94%) | 78 (98%) | 63 (98%) |
| Lives alone (No) | 24 (80%) | 24 (77%) | 65 (81%) | 47 (73%) |
| Carer gender (Female) | 21 (70%) | 22 (71%) | 60 (75%) | 48 (75%) |
| Carer age | 70 | 72 | 67 | 69 |

#### Appendix 3 - Supplementary tables for training and delivery costs

**Appendix 3, Table 1. Training costs for Two-Day Course (2 Trainers, 2 Therapist and 4 RSWs)**

| **Cost Categories for Training** | **In Person** | **Online** |
|---|---|---|
| Cost of two trainers (£195.17 per day per trainer x 2 trainers x 2 days) | £781 | £781 |
| Cost of mileage for 8 people (2 trainers + 6 therapists) (25 miles average roundtrip x £0.45 per mile) | £180 | NA |
| Cost of manuals and printing for 6 therapists over 2-day training (£12.50 per person x 6) | £75 | NA |
| Cost of 2 therapist attending 2-day training (£195.17 per day per therapist x 2 therapists x 2 days) | £781 | £781 |
| Cost of RSWs attending 2-day training (£122.82 per day per RSW x 4 RSWs x 2 days) | £983 | £983 |
| Cost of lunch and refreshments for 8 people (8 people x 2 days x £10 for lunch and refreshments) | £160 | NA |
| Cost of venue hire (£200 per day x 2 days) | £400 | NA |
| **Total costs to deliver 2-day training for 8 people (2 trainers + 6 therapists)** | **£3,360** | **£2,545** |
| **Total number of PrAISED therapists trained** | **61** | |
| **Total number of 2-day training courses required to train 60 therapists** | **10** | |
| **Total number of PrAISED patient participants** | **183** | |
| **Average cost of a 2-day training per PrAISED patient participant** | **£184** | **£139** |

**Appendix 3, Table 2. Delivery Costs for In-Person programme (n=61)**

| | **PrAISED** | **Usual Care** |
|---|---|---|
| Number of participants with early dementia receiving PrAISED intervention | 30 | 31 |
| Average number of sessions delivered per participant | 26 | 2 |
| Total number of sessions delivered | 780 | 62 |
| Average time for therapist /RSW to deliver a session (including travel) | 2.5 hours | 2.5 hours |
| Total number of hours delivering PrAISED | 1,950 hours | 155 hours |
| Average number of sessions delivered per participant by registered therapists | 8 (32%) | 1 (50%) |
| Average number of sessions delivered per participant by support workers | 18 (68%) | 1 (50%) |
| Total number of registered therapist hours | 624 hours | 77 hours |
| Total number of support worker hours | 1,326 hours | 78 hours |
| Average mileage to and from participants' homes (round trip) | 25 miles | 25 miles |
| Total mileage to and from participants' homes | 19,500 miles | 1,550 miles |
| Total cost of mileage to and from participants' homes (£0.45 per mile) | £8,775 | £698 |
| Total cost of registered therapists (£26.92/hour) | £16,798 | £2,073 |
| Total cost of support workers (£16.94/hour) | £22,462 | £1,321 |
| Equipment costs (£30 per participant) | £900 | £0 |
| Stationery costs (£5 per participant) | £150 | £0 |
| **Total delivery costs** | £49,085 | £4,092 |
| **Total delivery costs per participant** | £1,636 | £132 |
| **Total training costs per participant** | £184 | £0 |
| **Total costs per participant** | £1,820 | £132 |
| **Difference in total costs per participant between groups** | **£1,688** | |

**Appendix 3, Table 3. Delivery Costs for Blended programme (n=144)**

| | PrAISED | Usual Care |
|---|---|---|
| Number of participants with early dementia receiving PrAISED intervention | 80 | 64 |
| Average number of sessions delivered per participant | 26 | 2 |
| Total number of sessions delivered | 2,028 | 128 |
| Total number of sessions delivered in person | 1,399 (69%) | 88 (69%) |
| Total number of sessions delivered remote (video conference + teleconference) | 629 (31%) | 40 (31%) |
| Total number of sessions delivered video conferences | 102 (5%) | 6 (5%) |
| Total number of sessions delivered teleconferences | 527 (26%) | 33 (26%) |
| Average time for therapist /RSW to deliver a session in person (including travel) | 2.5 hours | 2.5 hours |
| Average time for therapist /RSW to deliver a session in video conferences | 1 hour | 1 hour |
| Average time for therapist /RSW to deliver a session in teleconferences | 0.5 hour | 0.5 hour |
| Total number of hours delivering in person | 3,498 hours | 220 hours |
| Total number of hours delivering video conference | 102 hours | 6 hours |
| Total number of hours delivering teleconference | 264 hours | 17 hours |
| Total number of hours delivering | 3,864 hours | 243 hours |
| Average number of sessions delivered per participant by registered therapists | 8 (32%) | 1 (50%) |
| Average number of sessions delivered per participant by support workers | 18 (68%) | 1 (50%) |
| Total number of registered therapist hours | 1,236 hours | 121 hours |
| Total number of support worker hours | 2,628 hours | 122 hours |
| Average mileage to and from participants' homes (round trip) | 25 miles | 25 miles |
| Total mileage to and from participants' homes | 34,975 miles | 2,200 miles |
| Total cost of mileage to and from participants' homes (£0.45 per mile) | £15,739 | £990 |
| Total cost of registered therapists (£26.92/hour) | £33,273 | £3,257 |
| Total cost of support workers (£16.94/hour) | £44,518 | £2,067 |
| Equipment costs (£30 per participant) | £2,400 | £0 |
| Stationery costs (£5 per participant) | £400 | £0 |
| **Total delivery costs** | £96,330 | £6,314 |
| **Total delivery costs per participant** | £1,204 | £99 |
| **Total training costs per participant** | £139 | £0 |
| **Total costs per participant** | £1,343 | £99 |
| **Difference in total costs per participant between groups** | **£1,244** | |

#### Appendix 4 - Supplementary tables for health service resource use

**Appendix 4, Table 1. Health Service Resource Use of In-person programme**

| **PrAISED Patients (n=30)**<br>**Usual Care Patients (n=30)** | **Baseline** | **12-mos** | **Quantity Difference** | **Cost per Visit** | **Total Cost** | **Average Cost per Patient** |
|---|---|---|---|---|---|---|
| GP visits (PrAISED) | 27 | 40 | 13 | £42/visit | £546 | £18.20 |
| GP visits (usual care) | 25 | 20 | -5 | £42/visit | -£210 | -£7.00 |
| Admiral nurse visits (PrAISED) | 1 | 0 | -1 | £41/visit | -£41 | -£1.37 |
| Admiral nurse visits (usual care) | 0 | 0 | 0 | £41/visit | £0 | £0.00 |
| Mental Health nurse visits (PrAISED) | 4 | 0 | -4 | £41/visit | -£164 | -£5.47 |
| Mental Health nurse visits (usual care) | 1 | 6 | 5 | £41/visit | £205 | £6.83 |
| District nurse visits (PrAISED) | 15 | 18 | 3 | £41/visit | £123 | £4.10 |
| District nurse visits (usual care) | 6 | 12 | 6 | £41/visit | £246 | £8.20 |
| Specialist nurse visits (PrAISED) | 8 | 2 | -6 | £51/visit | -£306 | -£10.20 |
| Specialist nurse visits (usual care) | 0 | 2 | 2 | £51/visit | £102 | £3.40 |
| Practice nurse visits (PrAISED) | 24 | 29 | 5 | £44/visit | £220 | £7.33 |
| Practice nurse visits (usual care) | 16 | 25 | 9 | £44/visit | £396 | £13.20 |
| Physiotherapist visits (PrAISED) | 5 | 6 | 1 | £69/visit | £69 | £2.30 |
| Physiotherapist visits (usual care) | 2 | 0 | -2 | £69/visit | -£138 | -£4.60 |
| Occupational therapist (PrAISED) | 0 | 4 | 4 | £87/visit | £348 | £11.60 |
| Occupational therapist (usual care) | 2 | 3 | 1 | £87/visit | £87 | £2.90 |
| Outpatient services (PrAISED) | 24 | 16 | -8 | £229/visit | -£1,832 | -£61.07 |
| Outpatient services (usual care) | 12 | 9 | -3 | £229/visit | -£687 | -£22.90 |
| Emergency department service (PrAISED) | 1 | 3 | 2 | £297/visit | £458 | £15.27 |
| Emergency department service (usual care) | 2 | 1 | -1 | £297/visit | -£229 | -£7.63 |
| Total (PrAISED) | | | | | | -£19.31 |
| Total (usual care) | | | | | | -£7.60 |
| **Difference between groups** | | | | | | **-£11.71** |

**Appendix 4, Table 2. Health Service Resource Use of Blended programme**

| PrAISED Patients (n=79)<br>Usual Care Patients (n=62) | Baseline | 12-mos | Quantity Difference | Cost per Visit | Total Cost | Average Cost per Patient |
|---|---|---|---|---|---|---|
| GP visits (PrAISED) | 71 | 45 | -26 | £42/visit | -£1,092 | -£13.82 |
| GP visits (usual care) | 72 | 55 | -17 | £42/visit | -£714 | -£11.52 |
| Admiral nurse visits (PrAISED) | 2 | 0 | -2 | £41/visit | -£82 | -£1.04 |
| Admiral nurse visits (usual care) | 0 | 0 | 0 | £41/visit | £0 | £0.00 |
| Mental Health nurse visits (PrAISED) | 7 | 6 | -1 | £41/visit | -£41 | -£0.52 |
| Mental Health nurse visits (usual care) | 2 | 0 | -2 | £41/visit | -£82 | -£1.32 |
| District nurse visits (PrAISED) | 21 | 40 | 19 | £41/visit | £779 | £9.86 |
| District nurse visits (usual care) | 47 | 33 | -14 | £41/visit | -£574 | -£9.26 |
| Specialist nurse visits (PrAISED) | 8 | 4 | -4 | £51/visit | -£204 | -£2.58 |
| Specialist nurse visits (usual care) | 12 | 5 | -7 | £51/visit | -£357 | -£5.76 |
| Practice nurse visits (PrAISED) | 60 | 54 | -6 | £44/visit | -£264 | -£3.34 |
| Practice nurse visits (usual care) | 81 | 45 | -36 | £44/visit | -£1,584 | -£25.55 |
| Physiotherapist visits (PrAISED) | 5 | 12 | 7 | £69/visit | £483 | £6.11 |
| Physiotherapist visits (usual care) | 21 | 24 | 3 | £69/visit | £207 | £3.34 |
| Occupational therapist (PrAISED) | 5 | 10 | 5 | £87/visit | £435 | £5.51 |
| Occupational therapist (usual care) | 11 | 18 | 7 | £87/visit | £609 | £9.82 |
| Outpatient services (PrAISED) | 41 | 48 | 7 | £229/visit | £1,603 | £20.29 |
| Outpatient services (usual care) | 48 | 57 | 9 | £229/visit | £2,061 | £33.24 |
| Emergency department service (PrAISED) | 8 | 12 | 4 | £297/visit | £1,188 | £15.04 |
| Emergency department service (usual care) | 3 | 7 | 4 | £297/visit | £1,188 | £19.16 |
| **Total (PrAISED)** | | | | | | **£35.51** |
| **Total (usual care)** | | | | | | **£12.15** |
| **Difference between groups** | | | | | | **£23.36** |

**Appendix 4, Table 3. Health service resource use cost reference.**

| Service | Unit Cost (£) | Reference |
|---|---|---|
| GPs | 42 | PSSRU (hospital-based health care staff: Foundation doctor FY2 40 hr/wk) |
| Admiral nurses | 41 | PSSRU (hospital-based health care staff: nurse band 5) |
| Mental Health nurses | 41 | PSSRU (hospital-based health care staff: mental health nurse band 5) |
| District nurses | 41 | PSSRU (hospital-based health care staff: nurses band 5) |
| Specialist nurse | 51 | PSSRU (hospital-based health care staff: nurse band 6) |
| Practice nurse (GP clinic) | 44 | PSSRU (community based health care staff (costs included qualifications) |
| Physiotherapist | 69 | NHS costs 2020/2021 |
| Occupational therapist | 87 | NHS costs 2020/2021 |
| Outpatient clinic | 229 | NHS costs 2020/2021 (Outpatient procedures) |
| Accident & Emergency | 297 | NHS unit cost 2020/2021 (Not admitted) |

Appendix 5 – Questionnaires

#### Falls Efficacy scale - International (FES-I) – Short

**Now we would like to ask some questions about how concerned you are about the possibility of falling. Please reply thinking about how you usually do the activity.**

**If you currently don't do the activity, please answer to show whether you think you would be concerned about falling IF you did the activity.**

*(give participant prompt card)*

**For each of the following activities, please choose a statement which is closest to your own opinion to show how concerned you are that you might fall if you did this activity.**

*(circle one answer for each question)*

| | Not at all concerned (1) | Somewhat concerned (2) | Fairly concerned (3) | Very concerned (4) |
|---|---|---|---|---|
| Getting dressed or undressed | 1 | 2 | 3 | 4 |
| Taking a bath or shower | 1 | 2 | 3 | 4 |
| Getting in or out of a chair | 1 | 2 | 3 | 4 |
| Going up or down stairs | 1 | 2 | 3 | 4 |
| Reaching for something above your head or on the ground | 1 | 2 | 3 | 4 |
| Walking up or down a slope | 1 | 2 | 3 | 4 |
| Going out to a social event (e.g. religious service, family gathering or club meeting) | 1 | 2 | 3 | 4 |

### EQ5D-5L – proxy

*The respondent can complete this questionnaire by hand. However, the researcher should administer this questionnaire if this is preferred.*

**For each domain, please choose** (tick) **one statement that describes your FRIEND / RELATIVE'S health TODAY**

**1.1 Mobility**

1 No problems in walking about
2 Slight problems in walking about
3 Moderate problems in walking about
4 Severe problems in walking about
5 Unable to walk about

**1.2 Self-care**

1 No problems washing or dressing him/herself
2 Slight problems washing or dressing him/herself
3 Moderate problems washing or dressing him/herself
4 Severe problems washing or dressing him/herself
5 Unable to wash or dress him/herself

**1.3 Usual activities (e.g. housework, leisure, family)**

1 No problems doing their usual activities

2 Slight problems doing their usual activities

3 Moderate problems doing their usual activities

4 Severe problems doing their usual activities.

5 Unable to do their usual activities

**1.4 Pain / Discomfort**

1 No pain or discomfort

2 Slight pain or discomfort

3 Moderate pain or discomfort

4 Severe pain or discomfort

5 Extreme pain or discomfort

**1.5 Anxiety / Depression**

1 Not anxious or depressed

2 Slightly anxious or depressed

3 Moderately anxious or depressed

4 Severely anxious or depressed

5 Extremely anxious or depressed

**We would like to know how good or bad your FRIEND / RELATIVE'S health is TODAY**.

This scale is numbered from 0 to 100.
100 means the best health you can imagine.
0 means the worst health you can imagine.

**Mark an X on the scale to indicate how your FRIEND / RELATIVE'S health is TODAY.**

**Now, please write the number you marked on the scale in the box below.**

Your friend/ relative's health today =

The best health you can imagine

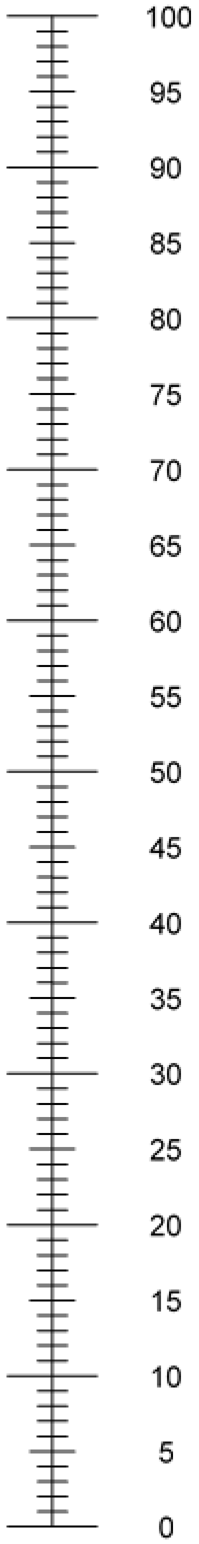

The worst health you can imagine

Carer Strain Index (CSI)

**These are questions about <u>you</u> and how you are feeling.**

**There is a list below of things which other people have found to be difficult when helping someone who has an illness. We would like to know if any of these apply to you over the last few weeks. Please answer all the questions by putting a tick in the box which you think most clearly applies to you.**

*Please tick one box for each item.*

| | |
|---|---|
| **1.1.** | **Sleep is disturbed** (e.g. because the person you care for is in and out of bed or wanders around at night)<br>☐ 1 Yes ☐ 0 No |
| **1.2.** | **It is inconvenient** (e.g. because helping takes so much time or it's a long drive over to help)<br>☐ 1 Yes ☐ 0 No |
| **1.3.** | **It is a physical strain** (e.g. because of lifting in and out of a chair; effort or concentration is required)<br>☐ 1 Yes ☐ 0 No |
| **1.4.** | **It is confining** (e.g. helping restricts free time or cannot go visiting)<br>☐ 1 Yes ☐ 0 No |
| **1.5.** | **There have been family adjustments** (e.g. because helping has disrupted my routine; there has been no privacy)<br>☐ 1 Yes ☐ 0 No |
| **1.6.** | **There have been changes in personal plans** (e.g. I had to turn down a job; could not go on vacation/holiday)<br>☐ 1 Yes ☐ 0 No |
| **1.7.** | **There have been other demands on my time** (e.g. from other family members)<br>☐ 1 Yes ☐ 0 No |
| **1.8.** | **There have been emotional adjustments** (e.g. because of severe arguments)<br>☐ 1 Yes ☐ 0 No |
| **1.9.** | **Some behaviour is upsetting** (e.g. because of incontinence; the person you care for has trouble remembering things; or the person you care for accuses people of taking things)<br>☐ 1 Yes ☐ 0 No |

| | |
|---|---|
| **1.10.** | **It is upsetting to find the person you care for has changed so much from his/her former self** (e.g. he/she is a different person than he/she used to be)<br>☐ 1 Yes ☐ 0 No |
| **1.11.** | **There have been work adjustments** (e.g. because of having to take time off)<br>☐ 1 Yes ☐ 0 No |
| **1.12.** | **It is a financial strain**<br>☐ 1 Yes ☐ 0 No |
| **1.13.** | **Feeling completely overwhelmed** (e.g. because of worry about the person you care for; concerns about how you will manage)<br>☐ 1 Yes ☐ 0 No |

Community activities questionnaire in follow-up questionnaire

1) During the past 3 months, has [your friend / relative] taken part in any organised group activities for at least once a month or more?

| | |
|---|---|
| | Cognitive Stimulation Therapy (CST) group |
| | Dance group |
| | Drama group |
| | Drawing/painting group |
| | Exercise group |
| | Memory clinic |
| | Pilates |
| | Reading group |
| | Swimming group |
| | Tai Chi |
| | Walking group |
| | Weight loss group |
| | Yoga group |
| | Other: please list any other community activities in boxes below |

2) Compared to 12 months ago, does [your friend / relative] participate in organised group activities?

| | |
|---|---|
| | Not at all |
| | Just a little |
| | Somewhat |
| | Moderately |
| | Quite a lot |
| | Very much |
